# Competency for Japanese Novice Medical Laboratory Scientists: A Delphi Method

**DOI:** 10.1101/2022.03.17.22272530

**Authors:** Kiriko Maekawa, Sayaka Kotera, Hiroyuki Ohsaki

## Abstract

**Introduction:** Competency is used to channel abilities into successful processes and is employed in the medical field. Worldwide, many laboratory competencies are available, but little evidence-based information about the competency of novice medical laboratory scientists exists in Japan. This study aimed to clarify competencies by various expert opinions around Japan.

**Methods:** The Delphi method was used to achieve an expert consensus on the competencies of novice medical laboratory scientists. We asked the participants to evaluate the importance of each item using the Likert scale and set 70% as the final consensus rate. The participants comprised medical laboratory scientists who met the following inclusion criteria: (1) qualified Japanese national medical laboratory scientists, (2) practised with >10 years of experience and (3) were heads of medical laboratory scientists in a clinical setting.

**Results:** We obtained 106 responses from 400 (26.5%) participants in round one and 95 from 106 (89.6%) participants in round two. Their professional experience mean ± standard deviation was 32.4 ± 6.0 years (range: 13–41). The average of each category consensus rate was >99.1%. The opinions of 95 experts converged, who agreed that competency comprised 8 categories and 54 items.

**Discussion:** Here, Japanese novice medical laboratory scientists required not only basic laboratory competencies but also other competencies, such as ‘medical safety management’, ‘interpersonal relationships’, ‘professional development’ and ‘ethics’, and some items were characteristic of Japan. Further research is warranted to explore assessments tools through the scale development of competencies, thus helping clarify the differences between ability and correlated factors.

## Introduction

In recent years, the quality of medical care in Japan has been changing because of the declining birth rate and ageing population, thus developing the medical environment and increasing the sophistication and specialisation of medical technology. These changes affect the work of doctors and nurses and several medical professionals, including a medical laboratory scientist (MLS)^*^. Specifically, in 2020, the spread of coronavirus disease 2019 (COVID-19) has led to an increased focus on polymerase chain reaction (PCR) and antibody testing, increasing the need for MLS [1]. To cope with these changes, training institutions for MLS have reviewed their educational content [2]. Recently, the Japanese Ministry of Health, Labour and Welfare (MHLW) has reviewed the school course content and the number of credits in the curriculum of training institutes for MLSs [3]; there has also been a move for aligning educational standards. Furthermore, the trends in MLS education have focused on the standardisation of educational content and clinical practice. However, there remains a lack of clear standardisation of education for novice MLSs who have obtained national qualifications and are employed in various medical institutions [4]. The time available for in-service training in the dynamic medical field is limited, and training mid-career and senior MLSs who can supervise in line with the abilities of each novice is difficult.

The Office of Personnel Management defines competency as ‘a measurable pattern of knowledge, skills, abilities, behaviours and other characteristics that an individual needs to perform work roles or occupational functions successfully’ [5]. Competency is also used to manage abilities into successful processes and is widely employed in human resource development [6]. The International Federation of Biomedical Laboratory Science (IFBLS) and the Centers for Disease Control and Prevention (CDC) have developed competencies for biomedical laboratory scientists and public health laboratory professionals [7, 8]. Moreover, seven pathology industry associations in Australia have developed competency-based standards for medical scientists working in diagnostic pathology settings [9]. The standards are the entry level of a scientist into this profession, reflecting a combination of qualifications, skills and the assumption of personal responsibilities and accountability [7]. The American Society of Clinical Pathologists and the Canadian Society for Medical Laboratory Science offer competency-based certification examinations [10, 11] that are useful in determining whether candidates have the skills and knowledge required to succeed as MLSs [12]. Thus, competency is regarded as essential to ensure the abilities of MLSs. Nonetheless, we could not find any studies related to Japanese novice MLS competencies on PubMed, published from the earliest date up to October 2019. Because the importance of competencies has been examined and utilised in work and examinations internationally, the development of competencies in Japan would effectively improve MLS quality.

The worldwide movement towards having medical laboratories accredited by the International Organisation for Standardisation (ISO) 15189 is another reason for enhancing competency [13]. This international standard specifies requirements for quality and competence in a medical laboratory [14] and relates in part to services for clinical physiology. Hence, competency is clearly essential and fundamental for MLSs.

This study aimed to identify the competencies of novice MLSs using the Delphi method. The Delphi method is an interactive process for collecting and distilling the anonymous judgements of experts using a series of data collection and analysis techniques interspersed with feedback [15], and the approach enhances reliability [16]. There is little evidence-based information about the novice competencies of MLSs in Japan, and it would be essential to gather various expert views. These competencies are the minimum requirement for MLSs to be licensed and perform their jobs and have the advantage of setting clear goals for them and providing their supervisors with a systematic and objective way of educating them. Additionally, our study is expected to be used for their primary education in training schools.

## Method

### Study design

This study used the Delphi method [17, 18, 19], which is broadly employed in medical and health fields [21, 22], a useful tool that looks for answers to normative questions in the education area [23] in order to arrive at an expert consensus on the competencies of novice MLSs. The Delphi method is flexible [24], and we utilised the modified Delphi method [25], generating a draft competency using a literature review and some expert interviews rather than an initial round of the Delphi method. Subsequently, we distributed the competency questionnaires to gather the opinions of experts in Japan by using the Delphi method. In the competency questionnaires, we defined a novice MLS as one with <3 years of after-graduation experience.

### Instrument development

We developed a draft questionnaire by reviewing overseas literature pieces and publications on competencies [e.g. 7, 8, 13]. First, a draft questionnaire was constructed and modified based on the IFBLS’s core competency [7] subjects for biomedical laboratory scientists working in a general laboratory setting. Some items were modified for appropriateness with the clinical laboratory setting and Japanese context. Second, it was compared with the competency of the CDC guidelines and the association of a public health laboratory [8] to confirm any significant differences in terms of concept or direction. Lastly, some competency was added n referring to ISO 15189 [13] and other related literature about Japanese MLS requirements. The final modification was conducted downward from the professional to novice level. The content and face validity of the draft were assessed with professors of MLS education and MLSs. Through all the processes, the draft questionnaire was approved to become the first round Delphi questionnaire.

### The questionnaire

In the questionnaire, we asked the participants to evaluate the importance of each competence using a 4-point Likert scale (4 = important, 3 = somewhat important, 2 = somewhat unimportant, 1 = unimportant). Further, our study defined consensus as a positive agreement if the participants chose either 3 or 4 and no consensus as a negative agreement if they chose either 1 or 2 [26]. The recommended consensus rate of the Delphi method ranges from 51% to 70%, with 70% indicating more prudence [17]. Our study set 70% as the final consensus rate for each item, with two criteria [25]. An item receiving 4 points by more than 70% participants was automatically approved unless the item obtained comments and suggestions from the participants. Conversely, if an item received 1 point by more than 70% participants, it was automatically rejected. The item, except for the two criteria, would be approached based on the results and comments of the previous questionnaire round. Based on the two criteria, the Delphi survey was planning to conduct three rounds of administering the questionnaire. The study was undertaken at the beginning of the COVID-19 pandemic from January to March 2020.

### Participants

The participants comprised experienced MLSs who work at hospitals in Japan. Generally, a hospital has one head of MLSs, and the head takes reasonability for training and educating novice MLSs. They met the following criteria: (1) held the national qualification for MLSs in Japan, (2) work experience as an MLS for more than 10 years and (3) heads of MLSs or equivalent in a clinical setting. Heads of MLSs manage their laboratory and have enough knowledge, techniques, coaching skills and MLS experiences. Further, the hospital itself should be registered with the MLHW and have more than 300 beds [27]. In December 2020, 1,001 hospitals met the criteria through Japan, and 400 were chosen randomly after stratified sampling. The stratified sampling was used to reduce the sampling bias of regions and scales of hospitals. Additionally, we targeted the final responses of at least 30 hospitals, and the productive performance of the Delphi method [21] from 400 hospitals was recommended. The survey questionnaires were distributed by post all over Japan by using the postal address the MLHW described on the website [27]. To maintain anonymity, all postal addresses and names of the hospitals were managed by identification numbers and employed in a self-addressed envelope without any participant information.

### Data gathering

The participants were given the first questionnaire, comprising 8 categories and 51 competencies on a 4-point Likert scale. Three free comment boxes were added in respect of their right to free opinion. The reasons for participant ratings of 2 or 1 and ideas about any other necessary competency items and opinions about the study were filled in the comment boxes. Regarding demographics, position, years of experience as an MLS and the number of beds and MLS in the participant hospital were queried. The second survey was administered to the first participants, and they received the second questionnaire analysed based on the first survey with descriptive statistics and the comment results summarised. It comprised 8 categories and 54 competencies on the same rate scale as well as reasons for ratings of 2 or 1.

### Ethical consideration

Ethics approval to undertake this study was provided by the ethical committee of the Kobe University Graduate School of Health Sciences (approval number 904). Written informed consent was obtained from all the participants, whose confidentiality was preserved.

## Results

### Demographics

Table 1 presents the response rates in the first and second rounds and the characteristics of the experts. A total of 106 responses were received from 400 participants (26.5%) in round one and 95 from 106 (89.6%) in round two. The experts’ years of experience mean ± SD was 32.4 ± 6.0 years (range: 13–41). The numbers of beds and MLSs they belong to were 499.8 ± 204.3 and 34.9 ±22.1, respectively. Some participants reported that the number of beds was less than that of the MLHW registration.

**Table 1.** Total number and characteristics of the participants

|  | 1st round | 2nd round |
| --- | --- | --- |
| **Total** |  |  |
| Collections/distributions | 106/400 | 95/106 |
| (Response rate) | (26.5%) | (89.6%) |
| **Characteristics** |  |  |
| **Work experience years n (%)** |  |  |
| 10–19 years | 4 (3.8) |  |
| 20–29 years | 20 (19.0) |  |
| ≥30 years | 81 (77.1) |  |
| Mean ± SD (n = 105) | 32.4 ± 6.0 |  |
|  | (Range: 13–41 years) |  |
| **Number of MLSs in their department** |  |  |
| Mean ± SD | 34.9 ± 22.1 |  |
|  | (Range: 5–122) |  |
| **Number of beds in their department** |  |  |
| Mean ± SD (n = 103) | 499.8 ± 204.3 |  |
|  | (Range: 204–1200) |  |

All the collected items were analysed.

The second survey continued the survey of the participants from the first survey.

### Round 1

In the first round, the consensus rate in the Delphi survey was above 89.6% for 106 participants in all 51 items. Table 2 presents the overall consensus rate of each category. The reasons why item responses were answered as negative agreements were primarily related to the competency of difficulty for the novice MLSs. The comment responses suggested as necessary competencies are indicated below. The ‘Preparation and analysis (general)’ category obtained comments about the competency of external quality assessment, while ‘Preparation and analysis (physiology)’ received comments about the competency of communication with patients. Additionally, ‘Medical safety management’ earned comments about the competency of a related disaster; ‘Interpersonal relationships’ obtained comments about the basic competency of reporting, informing and consulting about the task and ‘Medical ethics’ received comments about the competency of manner as a member of society.

**Table 2.** Overall novice MLS competency categories based on the level of importance

| Competency category | 1st round (n = 106) |  |  | 2nd round (n = 95) |  |  |
| --- | --- | --- | --- | --- | --- | --- |
| | Items | Mean ± | Consensus | Items | Mean ± | Consensus |
|  |  | SD | (%) |  | SD | (%) |
| 1. Sample collection | 2 | 3.9 ± 0.0 | 99.5 | 2 | 3.9 ± 0.0 | 99.5 |
| 2. Preparation and analysis<br>(general) | 16 | 3.7 ± 0.1 | 97.3 | 17 | 3.7 ± 0.2 | 99.4 |
| 3. Preparation and analysis<br>(specimens) | 7 | 3.8 ± 0.1 | 98.2 | 6 | 3.8 ± 0.1 | 100.0 |
| 4. Preparation and analysis<br>(physiology) | 3 | 3.9 ± 0.1 | 99.0 | 5 | 3.8 ± 0.1 | 99.8 |
| 5. Medical safety management | 6 | 3.6 ± 0.2 | 96.9 | 6 | 3.7 ± 0.1 | 99.1 |
| 6. Interpersonal relationships | 6 | 3.6 ± 0.2 | 96.4 | 6 | 3.7 ± 0.1 | 99.5 |
| 7. Professional development | 5 | 3.6 ± 0.1 | 96.6 | 5 | 3.6 ± 0.1 | 99.4 |
| 8. Ethics | 6 | 3.7 ± 0.1 | 98.1 | 7 | 3.7 ± 0.2 | 99.4 |

In the first round, there were primarily two patterns of the modifications of competency. One was a downward modification, which was adopted for items that had less than 70% rating under ‘important’ following the comments from the participants who were out of consensus. The other was adding new items and ideas to the first questionnaire following the comments from the participants who thought about necessary competencies other than the questionnaire items. Owing to the modification, the first questionnaire had 8 categories and 54 items.

### Round 2

In the second round, the consensus rate was above 97.9% for 95 participants in all 54 items, including the newly added items. The comments from the participants were almost the same as in the first round. As per the results, the questionnaire was modified partially, and this round had become the final one because of the high consensus rate in each category (Table 2). Finally, the competency item had 8 categories and 54 items (Table3).

**Table 3.**
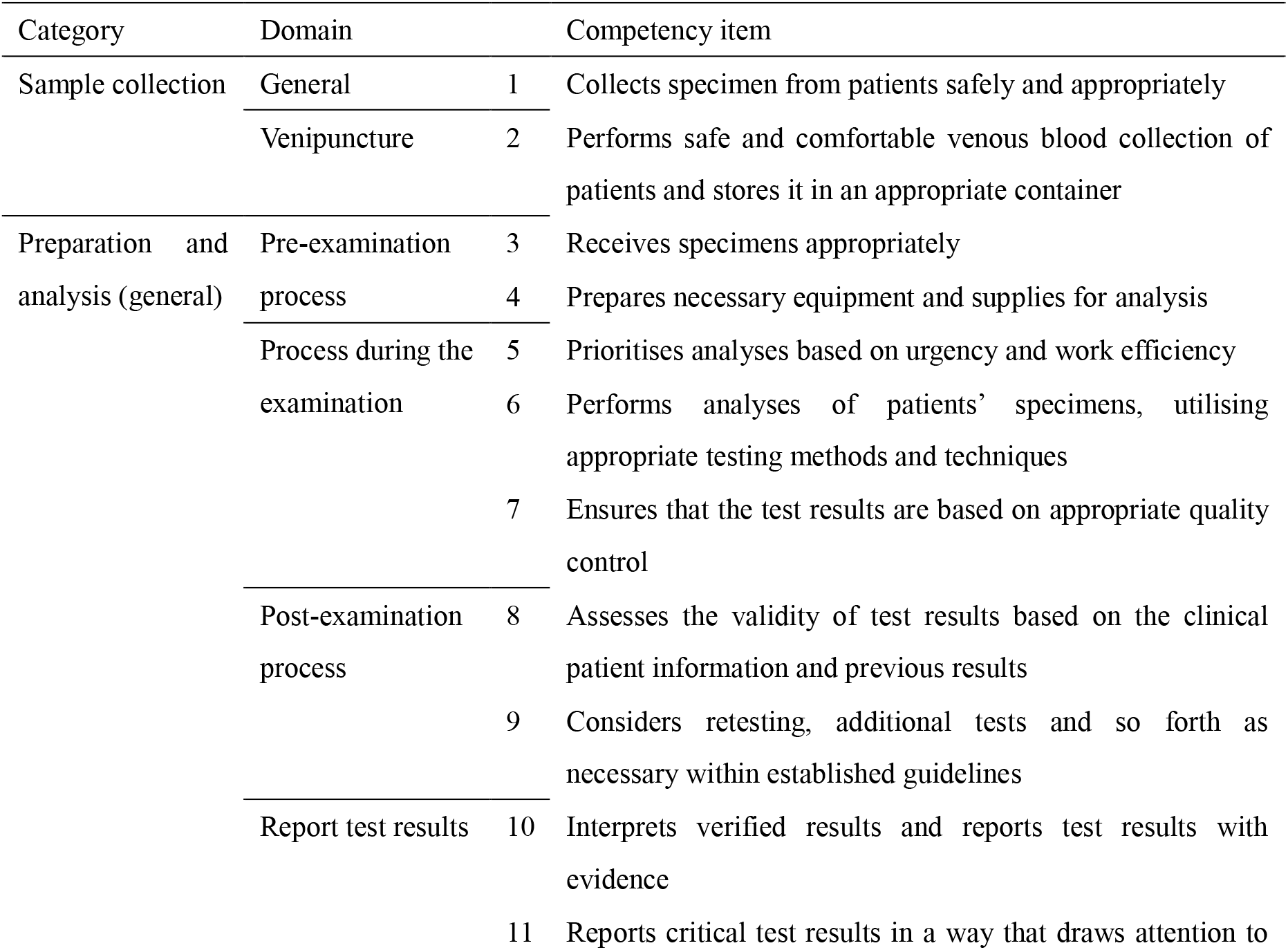
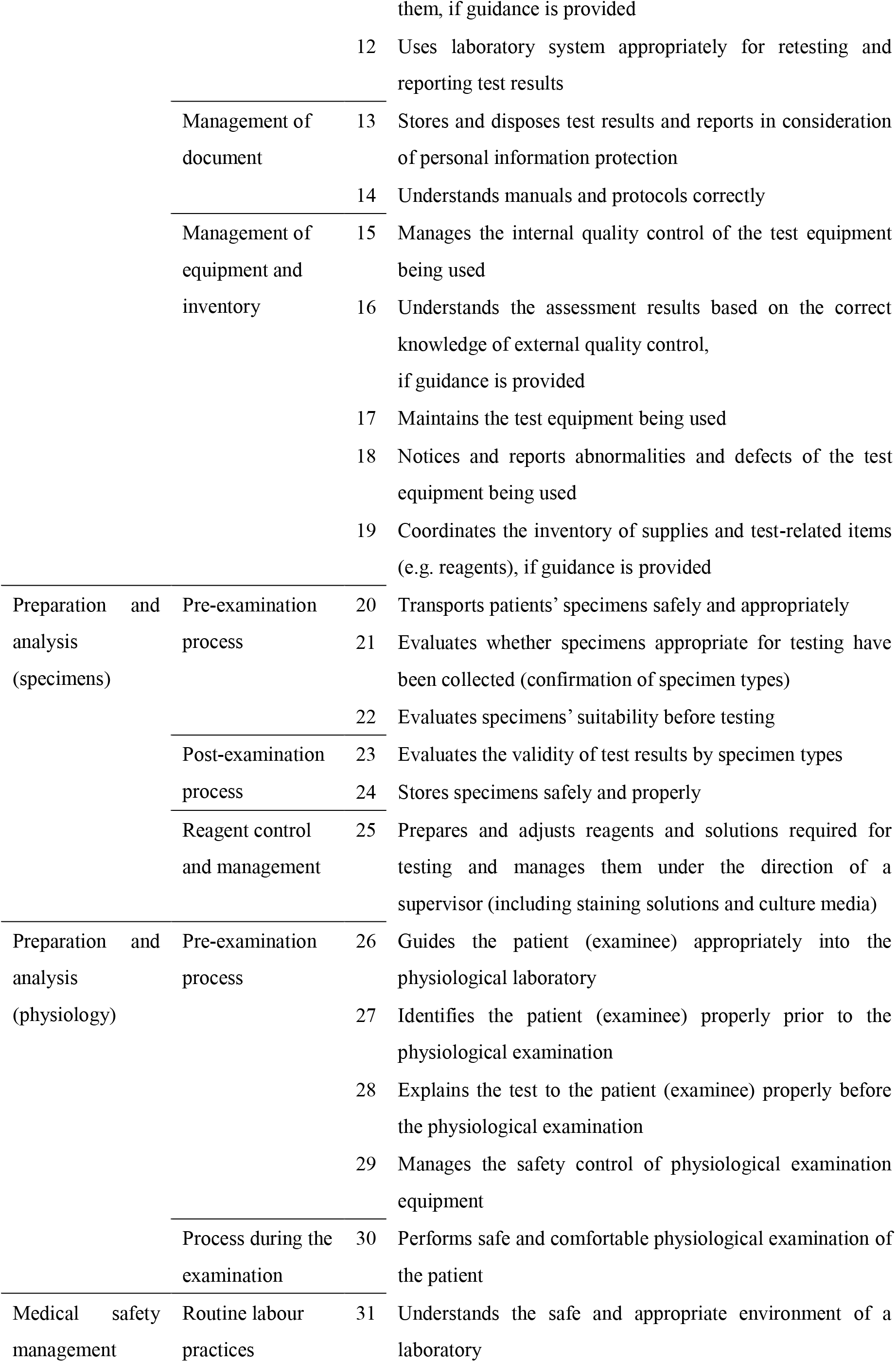
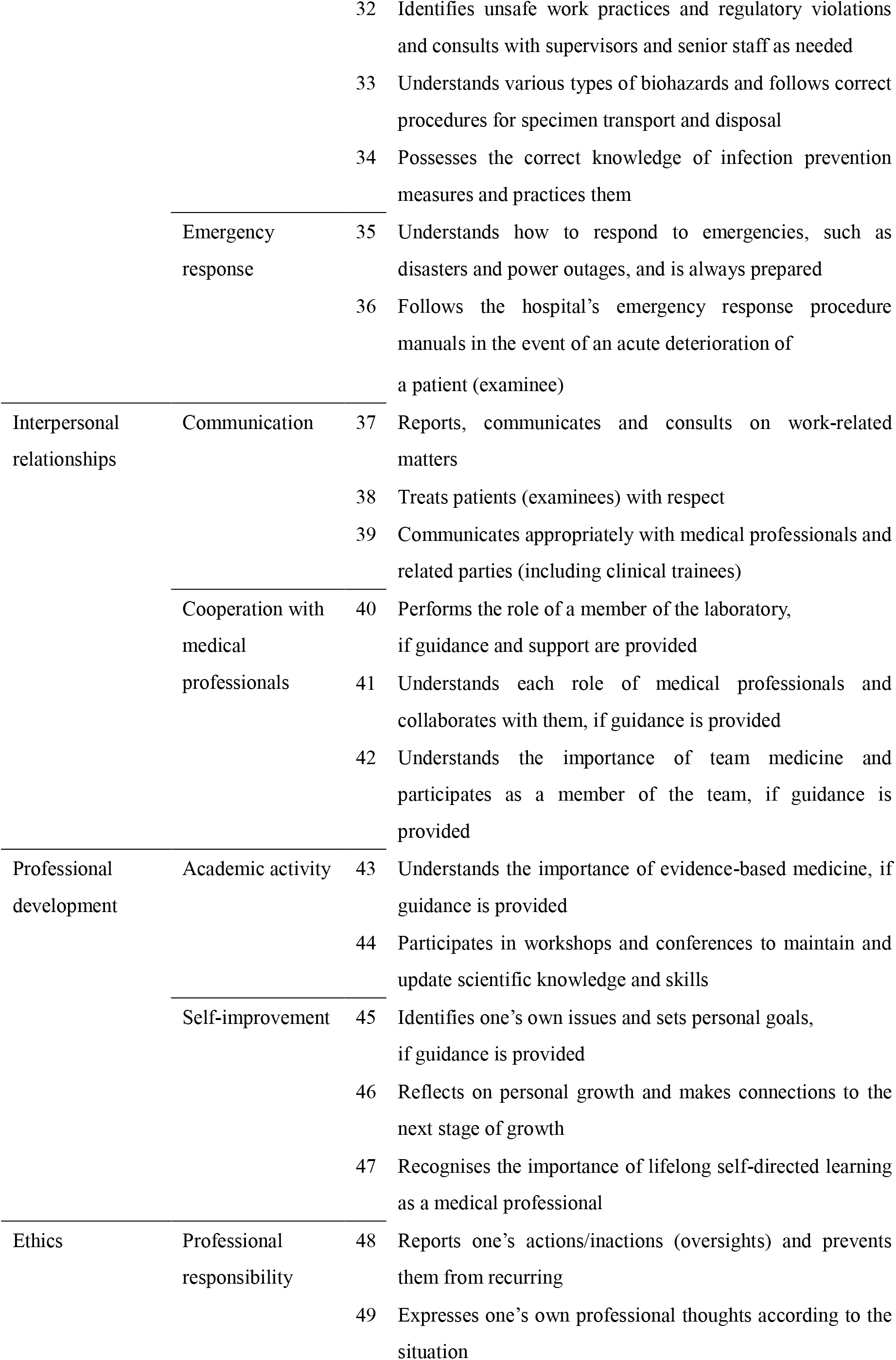
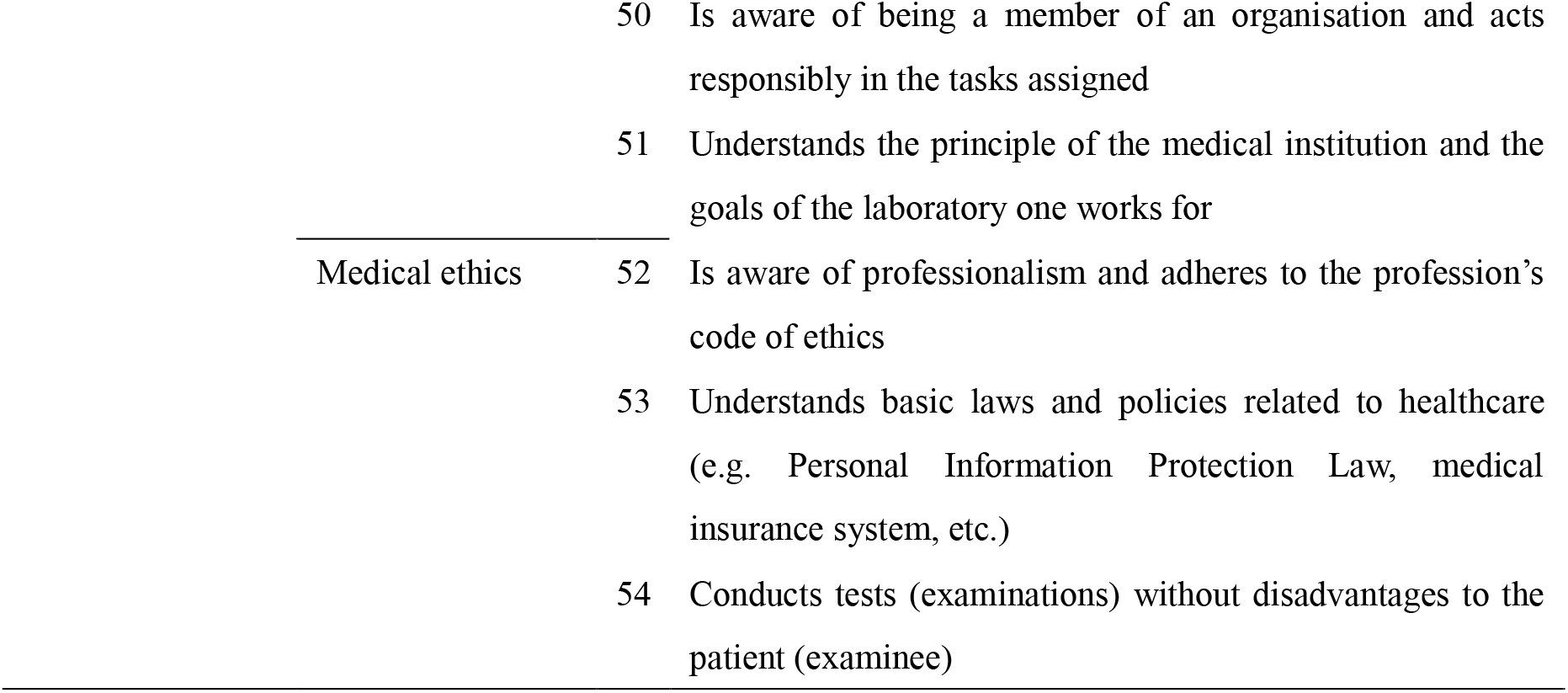
Japanese novice MLS competency in Japan

## Discussion

### Consensus rate

Our target for the final consensus rate, which was 70% and prudence, exceeded 97% in each category. The average consensus rate of each category was exceedingly high, and it was above 99.1%. The opinions of the 95 experts converged, and they agreed on the competency in this study.

### Findings

Several characteristics of competency are to be highlighted. The ‘Preparation and analysis (general)’ category was considered the ability to be more critical owing to the role expansion and increasing demand for PCR for COVID-19. The result clarified that the participants supported the importance of the categories. .Additionally, the ‘Preparation and analysis (physiology)’ category included examination and communication skills with the patients. One of the MLS roles is creating a comfortable environment where patients can take a physiological examination based on the quality of life [28]. Competency included ‘examination of patients with safety and comfort’ in such categories, supporting the role of MLS. Moreover, as per participant comments, ‘Medical safety management’ was added into the second survey questionnaire as a competency dealing with disaster situations. Japan is an earthquake-prone country, and disaster medicine is inevitable. Thus, MLS must accomplish medical testing tasks under disaster situations like a medical professional, and it is necessary to prepare responses for all contingencies in nonemergency periods [29]. As MLSs, the novices were recommended to understand the preparation required in developing competencies.

The survey results show that novice MLSs were expected to have a relatively higher ability in main laboratory skill competencies, such as the ‘Preparation and analysis’ category. Nevertheless, competencies in other categories required basic skills. A laboratory skill is considered essential, and minimum requirements as medical professionals were expected, such as a general responsibility as any worker in a clinical setting. The results showed high consensus rates across categories in the first round. By adding new items, the participants provided suggestions in the first round, which also had a high consensus rate in the second round.

Further research is warranted to explore the tools of assessments through the scale development of competencies, thus helping clarify the gaps of ability and correlated factors, which is also linked to study interventions.

Compared with the IFBLS competency, we reoffered to construct the draft questionnaire, and they were completely different in 2 categories (‘Sample collection’ and ‘Preparation and analysis (physiology)’ and 16 items. This difference was related to the divergent roles of MLSs, such as between Japan and other countries like the United States or European countries. Japanese MLSs get to collect some specimens and perform physiological laboratory work under the MLS qualification. However, MLSs in other countries do not, and instead of them, another professional does such work [30].

Another reason for some differences from the IFBLS competency was that our competency was limited to target workers in a healthcare institution and novice MLSs. The difference is pronounced in ‘interpersonal development skills’, where the IFBLS core competencies included interpersonal relationships with health professionals in other institutions, vendors and community members. Conversely, this study included only basic communication skills for working adults in healthcare institutions. Another one is the ‘Research ability’ category, which had two domains—’Academic activity’ and ‘Professional development’—in the first round. The IFBLS competencies required advanced abilities, such as research planning. Conversely, our competency weighted more reliable job performances than strengthening their research skills. Nonetheless, the participant comments revealed that developing an ability to acquire a high level of knowledge and skills through self-learning is required from the start of a new job. Therefore, we changed the category to ‘Professional development’ and the domains to ‘Academic activity’ and ‘Self-improvement’. The category competencies focused on self-study to enhance themselves.

### The panel

The demographics of the study participants were estimated to be a group of expert-level MLSs with a mean ± SD of 32.4 ± 6.0 years of experience. The expert opinion is placed at level 5 of the evidence level, and the Delphi method provides more substantial authority than the experts’ opinions [31]. Although the first response rate was low (26.5%), that of the second survey was high (95 of 106 [89.6%]), and the result suggested that the MLSs with a particular interest in in-service education participated in the study. The 95 participants fulfilled the requirement of the Delphi method [15]. The Delphi technique provided evidence of content and face validity, and the validity was influenced by the number of experts in the sample and the level of expertise and agreement [16]. This finding was suitable for those conditions, and it validated that competency had content and face validity. Thus, the mean ± SD number of MLSs in the participant departments was 34.9 ± 22.1, suggesting that the head of MLSs managed sufficient staff and had an in-service education. The mean ± SD number of beds in the facilities to which the participants belonged was 499.8 ± 204.3, approximately the same as the national average of 475.1 beds for facilities with more than 300 beds. The institutes for the participants were scattered all over the country. The participants in this study were professionally experienced head MLSs with a high level of awareness and interest in in-service education and who could educate their staff. Additionally, the results are free from bias because of the facility size and regional characteristics.

### Limitations

Our study has three limitations. First, the competencies were limited to MLSs working in medical institutions. It was a new challenge for Japanese MLS competencies, and we targeted MLSs in clinical settings. As the competencies required may differ according to another field of activity, such as public health laboratories, clarifying the competencies required in other institutions is essential.

Second, the participants were from clinical settings only, except for MLS educators at universities and colleges. Opinions from school and clinical educators for MLSs must be investigated. To summarise those opinions, MLS competencies should be widely utilised.

Third, the target institutions were limited to medical institutions with more than 300 beds, as they have sufficient numbers of MLSs, thus being able to ensure a proper educational system. Conversely, there may be difficulties in educating new appointees in small medical institutions with less than 300 beds. For instance, MLSs may not be available to spend enough time and effort on novice education because of the smaller number of personnel. Moreover, the scope of work may be broader than a large hospital, and duties with responsibility may be assigned at a relatively early stage.

This study revealed a tendency for new appointees to be required to have the ability to perform a wide range of laboratory tasks, and they were required for a relatively general responsibility. The study participants affirmed that the seniors and supervisors should mainly take up responsibilities during laboratory tasks, while the novices need primary responsibilities, such as the responsibility with regard to being members of an organisation and medical professionals. In the future, clarifying the actual conditions of work in small medical institutions and the issues involved in training novices will be crucial.

### Strengths and challenges

First, we believe that feedback regarding competencies can contribute to the development of in-service education. For supervisors, the competencies in this study provide the advantage that they can teach efficiently without dropping essential concepts. For novices, the clarification of the required competencies makes it easier for them to set their own goals and improve their work motivation. Although this study focused on the novice period, competency during this period can lead to competencies such as mid-career MLS and senior MLS competencies, and it is thus expected to develop an MLS career ladder. Given that the career ladder standardises and visualises the growth process of the required competencies [22, 23], it can be expected to develop systematic in-service education according to career levels.

Second, the novice competency can be utilised for MLS school education. The MHLW has been promoting the standardisation of education for MLSs [3], and it is thought to contribute to clinical practice guidelines. Additionally, this study is based on the core competencies of the IFBLS, and we believe that the inclusion of the competencies will pave the way for the development of world-class personnel.

Third, competencies could utilise research on MLS education. Compared with medical doctors, nurses and other medical professionals, MLSs have fewer education-based studies and information in Japan. This deviation may conceivably be because there are fewer people involved in research into the education of MLSs and because it is considered less important than others. In the future, MLS education will be enhanced by subsequent studies, which will support strengthening the identity and professionalism of MLSs. In this study, the Delphi method was utilised to identify the competencies as the minimum requirement for entry-level MLSs by the head of MLSs at a medical institution. The number of experts, level of expertise and agreement were sufficient to indicate the content and face validity of the competencies [16]. The expert group was willing to participate in the surveys, meaning that we could gather the opinions of those who are eager for clinical education. In the end, two surveys led to a convergence of keen expert opinions with a high consensus rate, which was more than 97.9%. The 54 competencies were identified for the 8 categories. The competencies were broadly consistent with those in other countries; however, some were unique to Japanese MLSs because Japanese MLSs handled the tasks of different professionals in other countries. Thus, we clarified that the requirements were specific to novice MLSs in Japanese medical institutions as a qualificator and professional MLS member.

## Data Availability

All data produced in the present study are available upon reasonable request to the authors

## Acknowledgement

This study would not be successful without the cooperation of the participants. The authors wish to thank all of the Delphi survey participants significantly despite the challenging situation owing to the COVID-19 pandemic. Additionally, we express our sincere thanks to the professors who provided valuable comments in the instrument development session.

## Declaration of interest

The authors declare no conflicts of interest.

## Footnotes

1 *A medical laboratory scientist (MLS), also known as clinical laboratory scientist (CLS), biomedical laboratory scientist (BLS) and medical technologist (MT), works in a medical laboratory.

## Notes

### Competing Interest Statement

The authors have declared no competing interest.

### Funding Statement

This study did not receive any funding

### Author Declarations

Ethics approval to undertake this study was provided by the ethical committee of the Kobe University Graduate School of Health Sciences (approval number 904).

